# LLM-Based Web Data Collection for Research Dataset Creation

**DOI:** 10.1101/2025.05.23.25328249

**Authors:** Thomas Berkane, Marie-Laure Charpignon, Maimuna Majumder

## Abstract

Researchers across many fields rely on web data to gain new insights and validate methods. However, assembling accurate and comprehensive datasets typically demands manual review of numerous web pages to identify and record only those data points relevant to specific research objectives. The vast and scattered nature of online information makes this process time-consuming and prone to human error. To address these challenges, we present a human-in-the-loop framework that automates web-scale data collection end-to-end using large language models (LLMs). Given a textual description of a target dataset, our framework (1) automatically formulates search engine queries, (2) navigates the web to identify relevant web pages, (3) extracts the data points of interest, and (4) performs quality control to produce a structured, research-ready dataset. Users remain in the loop throughout, able to inspect and adjust the framework’s decisions to ensure alignment with their needs. We introduce techniques to mitigate both search engine bias and LLM hallucinations during data extraction. Experiments across three diverse data collection tasks show our framework significantly outperforms existing methods, while a user-centered case study demonstrates its practical utility. We open-source our code to help other researchers create custom datasets more efficiently.

## 1 Introduction

Web data underpin research across many fields—including political science, economics, and public health—by enabling both method validation and new discoveries. Researchers typically depend on datasets previously collected by others, made publicly available by institutions, or assembled manually through web searches and information extraction. For example, news articles are valuable resources for reconstructing event timelines, tracking public attention, and compiling summary statistics. The data points contained in such articles become particularly useful when aggregated into datasets and validated prior to release. However, manual collection is time-consuming and risks missing relevant web pages or data points. For instance, the Johns Hopkins University COVID-19 dashboard (Dong et al., 2020), which reported global case and death counts, illustrates this challenge: data were initially collected manually from news and social media sources, but the process soon proved unsustainable and required automation.

In recent years, major social media platforms have restricted data access for both the public and researchers alike. Previously, social media data were often openly available through application programming interfaces (APIs), providing invaluable resources for studying socio-behavioral phenomena. Notable recent examples of access limitations include the shutdown of Meta’s CrowdTangle platform in August 2024 (Meta, 2024) and severe restrictions to Twitter’s (Coalition for Independent Technology Research, 2023) and Reddit’s APIs (Reddit Admin, 2023) in January and April 2023, respectively. These limitations have increased the need for researchers to independently collect data, often under significant time and budget constraints. To address these challenges, we present a framework for end-to-end, web-scale data collection that leverages recent advances in large language models (LLMs). Our method begins with a user-provided textual description of a target dataset, searches for relevant web pages and PDF documents, selectively extracts data, and produces a structured dataset. The framework is designed for human-in-the-loop interaction, allowing users to continuously monitor and adjust the data collection process to ensure alignment with their objectives. It maintains transparency by making each extracted data point traceable to its source via direct quotation. Further, we implement corrections for recency and geographical biases introduced by search engines and mitigate LLM hallucinations through source grounding. A final quality control step automatically flags potentially anomalous data points, such as outliers and duplicates, for manual review.

We evaluate the effectiveness of our framework on three diverse data collection tasks, for each of which we construct reference datasets via manual search and aggregation of results from multiple methods. We benchmark our framework against state-of-the-art tools, including ChatGPT-4o Deep Research (OpenAI, 2025) and Perplexity Deep Research (Perplexity, 2024), and observe substantial performance gains: for example, on the first data collection task, our framework improves F1-score by 74.3 and 38.5 percentage points compared to ChatGPT Deep Research and Perplexity Deep Research, respectively. Further, we conduct an ablation study demonstrating the effectiveness of our quality control step as well as our bias and hallucination mitigation techniques. Finally, a user-centered case study illustrates the practical value of our framework, showcasing both its strengths and real-world limitations.

## 2 Background and Related Work

### 2.1 Web Data Collection for Research

Most web data used in research consists of textual content from web pages, including news articles, social media posts, and PDF documents. Traditionally, researchers collect this data by visiting websites and manually recording relevant information—a labor-intensive and error-prone process. To improve efficiency, semi-automated methods relying on web scraping have emerged (Luscombe et al., 2022; Landers et al., 2016). However, these approaches require prior knowledge of website structures, limiting their generalizability across diverse and constantly changing web sources.

Many repositories aggregate online content, including those maintained by academic consortia, governments, and commercial platforms. Examples include Media Cloud (Roberts et al., 2021) and Google News^1^ for news, HealthMap (Freifeld et al., 2008) for public health, LexisNexis^2^ for legal data, and Federal Reserve Economic Data^3^ for economic statistics. While these platforms offer curated content and search capabilities, they are often domain-specific and may be proprietary.

In contrast, our approach provides an openaccess framework enabling researchers—including those without technical expertise—to collect and extract data from any web page or PDF document, across domains and use cases. This independence from data providers supports research objectivity and flexibility. The framework’s ability to quickly gather up-to-date web data empowers researchers to study emerging phenomena—such as disease outbreaks, climate events, or social crises—before conventional datasets are released.

### 2.2 Large Language Models

Recent advances in large language models (LLMs) have transformed web search and information access. Platforms such as Perplexity^4^ and ChatGPT Search^5^ can now answer user queries using up-to-date web information, extending LLM knowledge beyond their original pretraining data. More recently, Deep Research functionalities offered by LLM providers (Perplexity, 2024; OpenAI, 2025) have combined advanced LLM reasoning with active web navigation, enabling synthesis of detailed reports after extended web exploration.

LLMs have also been used to augment search processes in various ways. For example, they can reformulate queries to improve search effectiveness (Dhole and Agichtein, 2024) or generate query variants to expand coverage (Alaofi et al., 2023). In information retrieval, LLM-powered re-ranking models can supplement traditional search engines by computing semantic relevance scores for retrieved documents (Nogueira and Cho, 2020; Shakir et al., 2024), thus combining broad coverage with precise matching.

Beyond retrieval, LLMs have shown strong capabilities in data extraction (Polak and Morgan, 2024). Projects such as Crawl4AI^6^ leverage LLMs to extract structured information from unstructured web content. However, such approaches remain susceptible to hallucination—the generation of inaccurate or fabricated content presented as fact (Huang et al., 2025). Moreover, most existing tools focus on extraction alone, rather than supporting full end-to-end dataset creation. Our framework addresses these challenges by grounding extractions with source citations (Colverd et al., 2023) and highlighting specific text spans (Gero et al., 2023) for verification. This approach improves both reliability and transparency of research data.

## 3 Methodology

Figure 1 shows the steps of our framework.

**Figure 1:**
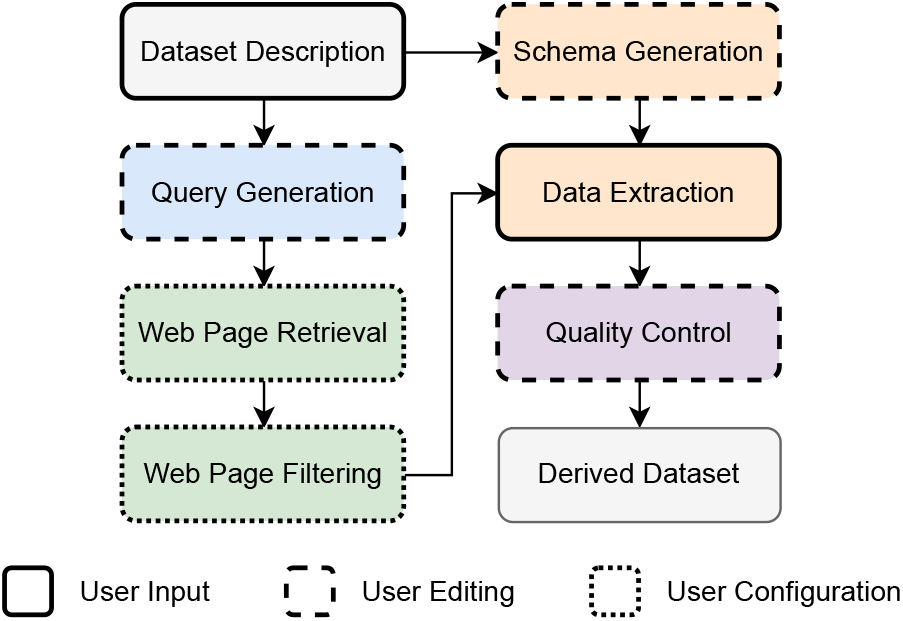
Overview of our LLM-based framework for web-scale data collection and research dataset creation. Users interact with our framework in three ways: providing instructions, editing intermediate results generated by the framework, and configuring parameters.

### 3.1 Search Query Generation

To enable the creation of both longitudinal and cross-sectional datasets—often requiring many similar searches (e.g., retrieving the same information for multiple countries or years)—our framework adopts a template-based query system. This enables efficient generation of structurally similar queries through placeholder substitution.

Query generation consists of two main components: templates and values. Templates define search query structures containing placeholders (denoted by curly braces {}, as shown in Figure 2) for variable elements. Values are the specific terms that replace these placeholders, provided by the user via a CSV file: placeholder names are listed in the header, and substitution values appear in subsequent rows. For templates containing multiple placeholders, the system produces queries for all possible value combinations. Figure 2 illustrates this template-based query generation process.

**Figure 2:**
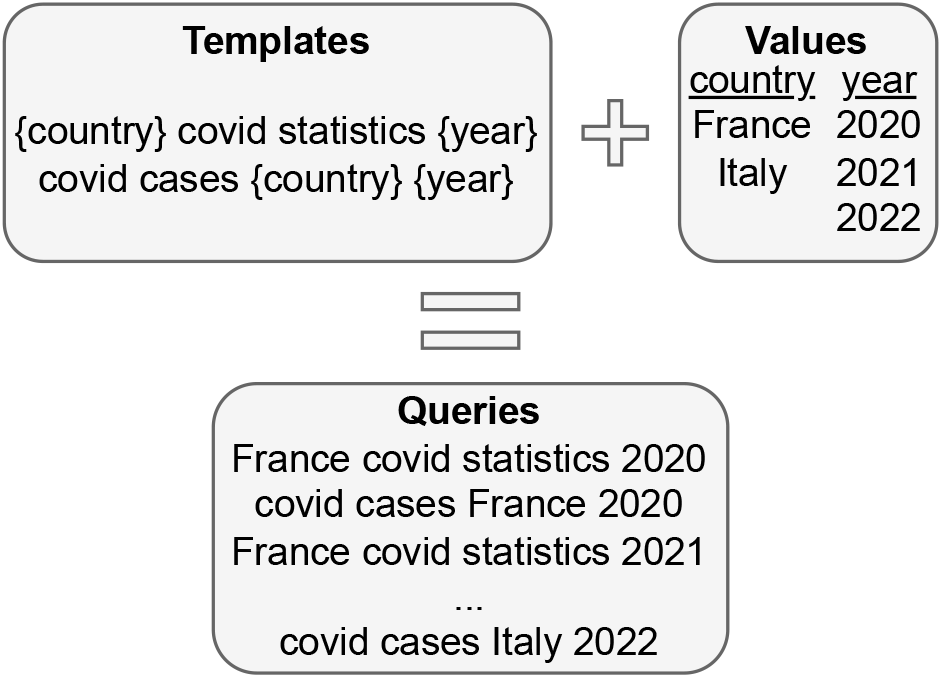
Illustration of our template-based search query system, showing how it combines search query templates with user-defined values to generate a large set of final search queries.

Our method requires two user inputs: a natural language description of the target dataset and a CSV file containing placeholder names (as headers) and corresponding values (as rows). The system submits a prompt—comprising the dataset description and CSV header—to Anthropic’s Claude 3.7 Sonnet model API (Anthropic, 2025) (see Appendix A). The model returns template-based Google Search queries intended to retrieve web pages containing relevant data. Users can then review and adjust the generated templates to ensure they meet their data collection objectives.

### 3.2 Web Page Retrieval

Once the user has validated the generated query templates, our framework combines them with the provided values to produce the final set of queries, which it then executes via Google Search. For each template-value combination, our system scrapes a user-defined number of search results pages—including web page titles and URLs—using Selenium (Selenium Project, 2024). By default, we set the maximum number of results pages to scrape to five. We determine this value empirically by noticing that the benefit of additional results pages diminishes rapidly. It can be either decreased to derive a structured dataset faster or increased to derive a more complete dataset. Our framework then removes duplicate URLs.

The retrieval process includes an optional date range filtering functionality, which uses Google Search’s before: and after: operators to temporally constrain search results. When enabled, searches are run both with and without this filter to ensure coverage of pages lacking metadata dates.

Google Search rankings are influenced by factors such as relevance, search engine optimization, site authority, and personalization. As a result, the top-ranked pages may not always be the ones that contain the target data. To address this, we introduce a re-ranking step, employing a pre-existing re-ranking model (Shakir et al., 2024) (Apache 2.0 license) to evaluate the relevance between each web page title and the query with which it was retrieved. This model assigns a relevance score (0 to 1) to each page. We retain pages only if they exceed a user-adjustable threshold (default 0.2, empirically found to balance inclusiveness and relevance); increasing the threshold restricts retention to more relevant pages, while decreasing it captures potentially more data points.

To expand data collection, we use snowball sampling, following hyperlinks on initially retrieved relevant web pages. This is particularly useful for discovering PDF documents, which are frequently linked within web pages and often contain valuable data. Using the Crawl4AI Python package, we extract all hyperlinks from relevant web pages and apply the re-ranking model to evaluate their relevance to the original query, adding those above the threshold to our set of relevant web pages. This process can, in principle, be repeated iteratively to expand the pool of distinct data sources.

### 3.3 Data Extraction

Our framework first generates a structured Pydantic (Colvin et al., 2025) schema by prompting Claude 3.7 Sonnet (see Appendix B), using the initial dataset description and CSV header. This schema specifies the fields to be extracted for each data point, including (i) one field per placeholder name in the CSV header to track each data point’s categories, (ii) data fields inferred from the dataset description, (iii) a date field, and (iv) a grounding text span—a short snippet from the source text where the data point was extracted—to facilitate user auditing and help mitigate LLM hallucinations and assist quality control (see Section 3.4).

For example, if the CSV header contains “country” and “year,” and the dataset description is “I want to build a dataset of covid case counts,” the schema would include fields for “country,” “year” (from the header), “case_count” (inferred by Claude), and the default “date” and “grounding_text_span” fields.

We extract text from web pages using Crawl4AI, which converts the content of each page to markdown, from which we remove hyperlinks. We convert PDFs to markdown via Mistral OCR (Mistral AI Team, 2025). Next, we prune markdown content to eliminate irrelevant sections (e.g., headers, navigation, unrelated text), thereby reducing the amount processed by Claude. The pruning process segments text into chunks, truncates large chunks, and re-ranks them by relevance to the original search query, keeping those above the 90th percentile—an empirically selected threshold that users may adjust. To ensure key temporal information (such as publication dates that are often pruned because they appear in isolated chunks on web pages) is retained, we also preserve any chunk with a relevance score with respect to the keyword “date” above 0.2, regardless of its query relevance. Finally, our framework applies Crawl4AI with Claude 3.7 Sonnet to each pruned markdown-converted web page, extracting all relevant data points and structuring them as JSON according to the generated schema. Users may optionally supply additional extraction instructions (e.g., “Only extract earthquake events after 2020”).

### 3.4 Quality Control

To mitigate potential hallucinations by the Claude model, we retain only data points whose grounding text is present in the original web page text. We then prompt Claude to conduct a sanity check of the assembled dataset (see Appendix C), presenting the complete dataset in CSV format for review. Claude inspects each data point individually, flagging potential issues for user review. While Claude does not resolve these issues, flagging suspicious entries helps accelerate manual data cleaning. This quality control step acts as a safeguard against extraction errors and ensures overall dataset coherence, facilitating identification of potential outliers, duplicates, or trends inconsistent with the dataset’s expected structure (e.g., non-monotonic time series for cumulative case counts).

## 4 Bias Mitigation

### 4.1 Recency Bias

Google Search results exhibit significant recency bias: web pages updated more recently are typically ranked higher. While this bias can help for certain applications such as monitoring current events, it is undesirable when building event timelines or longitudinal datasets, for example.

To counteract this, we introduce an optional time-chunking functionality when retrieving web pages over a date range spanning more than one year. This approach divides the date range into equal-size intervals of at most one year each—a maximum determined empirically, as retrieval rates remain relatively stable within one year but start to drop off beyond that. For each interval, we use Google Search’s before: and after: operators to collect web pages, thereby achieving a more uniform temporal distribution.

### 4.2 Geographical Bias

Personalization in Google Search also introduces geographical bias, with results varying based on the user’s location. Running queries as though they originate from the country of interest yields different source distributions—favoring local news, international organizations, humanitarian sites, and scientific publications, and reducing the prominence of English-language outlets and social media. This often improves both the quality and relevance of results, especially for queries about local events.

To take advantage of this, our framework lets users specify a country for each query. This sets the gl (geolocation) parameter in the Google Search query, instructing Google to return results as if the search originated from that country. If no country is specified, we default to the user’s current location.

## 5 Experiments

### 5.1 Baselines

We compare our framework to Perplexity Deep Research and ChatGPT-4o Deep Research. We do not include manual human web search as a baseline, as a fair comparison would require rigorous protocols to control for variability in human skill and effort, which is beyond the scope of this work.

### 5.2 Tasks

To evaluate our framework, we conduct experiments on three web data collection tasks selected for their topical diversity and the absence of central repositories for these data: (1) COVID-19 contact tracing app usage in the U.S.; (2) timelines of climate events in Haiti and Cameroon; and (3) characterization of police misconduct cases in Tennessee (see Table 1). For each task, we build a reference dataset by considering the union of all manually validated data points identified by four methods: Perplexity Deep Research, ChatGPT Deep Research, our framework, and manual web human search. Subsequently, we evaluate the extent to which the output of each evaluated method overlaps with this baseline. By aggregating outputs from these four methods, we ensure that any data point discovered by any approach is included in the reference dataset. While this does not guarantee complete coverage of all relevant data available on the web, it maximizes coverage within our evaluation. Further, we remove duplicates and entries with missing fields (e.g., an event without the date at which it occurred) to ensure reference dataset quality.

**Table 1:**
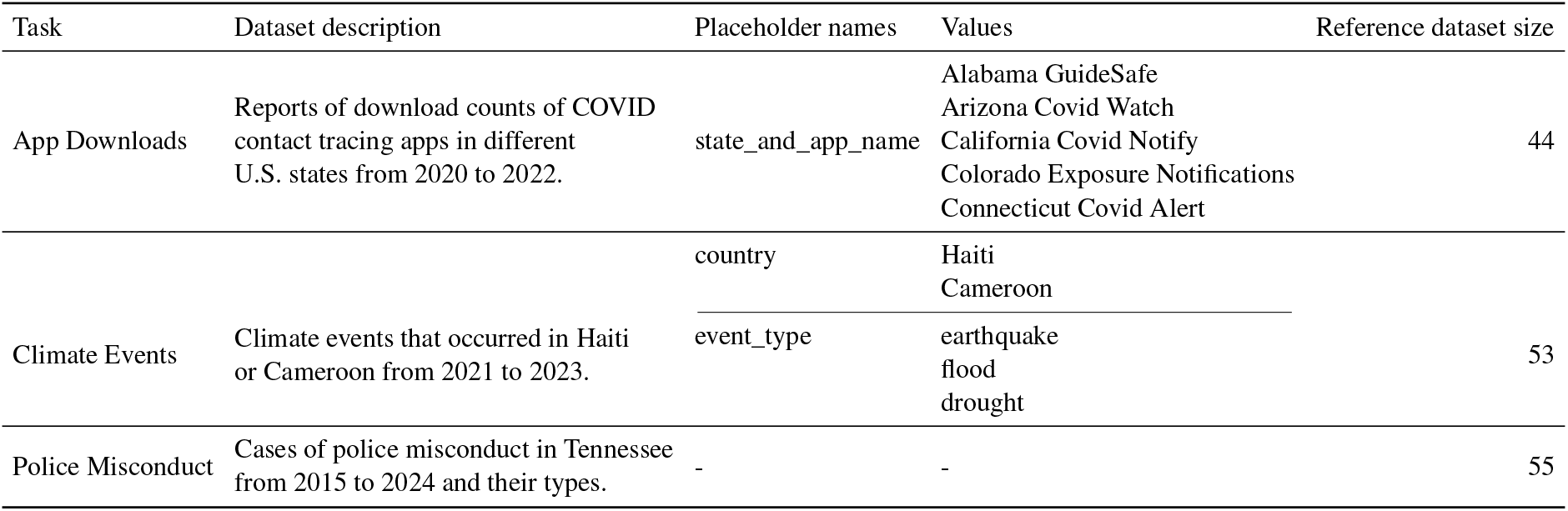
Summary of the three web data collection tasks used for evaluation. “Reference dataset size” indicates the number of manually validated data points against which we evaluate each method’s output. For the app downloads task, we only consider the first five states in alphabetical order due to limited computing resources.

### 5.3 Evaluation Protocol

Each method is run once per task, using the same dataset descriptions and placeholder names/values (Table 1) to avoid prompt tuning bias. Baselines use the prompt format in Appendix D. We provide details on the parameter settings for our framework in Appendix 4. While our framework produces structured data directly, Perplexity and ChatGPT Deep Research output free-form reports, which we manually parse to extract structured data points.

Because web data collection is inherently openended, extracted data points may not exactly match those in the reference dataset but can still be considered correct (e.g., if reported by a different outlet or on a nearby date). Since each reference dataset contains fewer than 100 entries, we manually compare each data point produced by any method to the reference dataset, applying task-specific rules to determine correctness. The matching window for dates is domain-specific, reflecting the typical temporal granularity of each data type (e.g., app download counts are reported approximately once a week, while droughts typically span months). More specifically, for the app downloads task, we match data points if they report the same state and download number within a one-week window. For climate events, we match earthquakes by exact country and day, floods by country and dates within one week, and droughts by country and year. We match police misconduct cases if they refer to the same incident (i.e., same officer, time, and type of misconduct). We leave *automated* (as opposed to *manual*) rule inference and matching of collected data against the reference dataset for future work.

We consider a data point correct if it matches a reference dataset entry according to these rules. For our framework, we also count a data point as correct if it is flagged by the quality control step as potentially problematic but can be easily fixed by a user. To ensure a fair comparison, we apply the same standard to the baselines: a data point is counted as correct if any easily resolvable issue is present—such as a missing date that can be recovered by visiting the source web page—but not if the issue involves incorrect or hallucinated content, such as an erroneous date. Data points that do not match any reference dataset entry are considered incorrect. Additionally, for our framework, we count any mis-extracted data point that is not flagged by quality control as incorrect. Before evaluation, we remove duplicate data points and exclude any data that fall outside the scope of the task, such as those outside the specified locations or date range.

For each method and task, we compute recall, precision, and F1-score.

## 6 Results and Analysis

Table 2 presents the results of our experiments. Across all three data collection tasks, our framework consistently achieves the highest recall and F1-score, with ChatGPT Deep Research performing second best, and Perplexity Deep Research ranking third. The performance gap between methods is substantial. However, precision varies by task, with no single method achieving the highest precision across all tasks. All methods are susceptible to extraction errors, with incorrect dates representing the most common mistake.

**Table 2:** Comparison of our approach against baselines across our three web data collection tasks.

| Method | Recall | Precision | F1-score |
| --- | --- | --- | --- |
| **APP DOWNLOADS** |  |  |  |
| Perplexity | 6.8 | 75.0 | 12.5 |
| ChatGPT | 31.8 | **100.0** | 48.3 |
| Ours | **81.8** | 92.3 | **86.8** |
| **CLIMATE EVENTS** |  |  |  |
| Perplexity | 11.3 | **100.0** | 20.3 |
| ChatGPT | 30.2 | 88.9 | 45.1 |
| Ours | **73.6** | 75.0 | **74.3** |
| **POLICE MISCONDUCT** |  |  |  |
| Perplexity | 12.7 | **100.0** | 22.6 |
| ChatGPT | 30.9 | **100.0** | 47.2 |
| Ours | **78.2** | 93.5 | **85.2** |

In the climate events task, our framework’s precision is comparatively lower because several earthquake events were incorrectly attributed to Cameroon. These errors stemmed from a source website that erroneously listed earthquakes as occurring in Cameroon, which our framework did not flag. Manual review later identified these as misclassifications, highlighting our method’s sensitivity to misleading sources and reinforcing the need for human validation of extracted data.

Unlike the fully autonomous deep research baselines, our framework is designed for human oversight throughout the data collection process—including duplicate removal and the resolution of flagged issues. This additional user input affords greater control and typically yields higher-quality datasets that are better aligned with research objectives. Because our system casts a wider net, it also retrieves more duplicate data points, especially for major events. While these duplicates can facilitate cross-source corroboration, they also increase the need for post-processing. By contrast, the deep research baselines retrieve fewer duplicates, reducing manual curation but still requiring user review to identify occasional errors or hallucinations.

The primary cost of our framework comes from using the Anthropic API to extract data from web pages; other steps—such as query and schema generation, PDF-to-markdown conversion, and quality control—incur negligible expenses. On average, extracting data from a single web page costs approximately $0.023 using our framework, with this cost remaining consistent across tasks.

### 6.1 Ablation Study

To assess the contribution of the quality control and bias mitigation components of our framework, we conduct an ablation study evaluating the effects of removing (i) source grounding, (ii) quality control (i.e., Claude flagging suspicious data points), (iii) recency bias mitigation (i.e., time-chunking), and (iv) geographical bias mitigation. For each variant, we disable a single component and measure the resulting change in performance across the three data collection tasks. Results are shown in Table 3.

**Table 3:** Ablation study results showing the impact of removing each component from our framework across the three web data collection tasks.

| Variant | Recall | Precision | F1-score |
| --- | --- | --- | --- |
| **APP DOWNLOADS** |  |  |  |
| Full framework | **81.8** | **92.3** | **86.7** |
| - Grounding | **81.8** | **92.3** | **86.7** |
| - Quality control | 77.3 | 87.2 | 82.0 |
| - Recency bias mitig. | **81.8** | **92.3** | **86.7** |
| - Geo bias mitigation | **81.8** | **92.3** | **86.7** |
| **CLIMATE EVENTS** |  |  |  |
| Full framework | **73.6** | **75.0** | **74.3** |
| - Grounding | **73.6** | 73.6 | 73.6 |
| - Quality control | 69.8 | 71.6 | 70.7 |
| - Recency bias mitig. | **73.6** | **75.0** | **74.3** |
| - Geo bias mitigation | 67.9 | 73.5 | 70.6 |
| **POLICE MISCONDUCT** |  |  |  |
| Full framework | **78.2** | **93.5** | **85.2** |
| - Grounding | **78.2** | **93.5** | **85.2** |
| - Quality control | **78.2** | 91.5 | 84.3 |
| - Recency bias mitig. | 14.0 | 80.0 | 23.8 |
| - Geo bias mitigation | **78.2** | **93.5** | **85.2** |

We observe that (i) removing source grounding has a slight negative impact only on the climate events task, where grounding enabled filtering of a hallucinated data point; for the other tasks, this ablation had no effect. While Claude 3.7 Sonnet rarely hallucinates, we expect source grounding to be more valuable when using lower-cost LLMs that may hallucinate more frequently, providing an important safeguard for data validity. (ii) Disabling quality control consistently reduces performance across all tasks, as this step helps flag problematic data points for user review and correction. (iii) Omitting recency bias mitigation only affects the police misconduct task, which spans a 10-year period; indeed, without time-chunking, Google Search retrieves far fewer relevant historical pages, highlighting the necessity of this step for long-term datasets. (iv) Removing geographical bias mitigation (i.e., always querying as if from the U.S.) negatively impacts only the climate events task, since the other two tasks are U.S.-specific; for climate events, geolocation allows our framework to retrieve more relevant sources local to Cameroon and Haiti.

Overall, these results highlight how each quality control and bias mitigation mechanism in our framework provides value for different types of data collection tasks.

### 6.2 User-Centered Case Study

We supplement our quantitative evaluation with a user-centered case study on the app downloads task, in which we examine both interaction with the human-in-the-loop components and whether the resulting dataset meets research needs. We walk through a typical data collection workflow, highlighting practical strengths and limitations from the user’s perspective.

We recruit a PhD-level public health researcher and assign them the following task: “During the COVID-19 pandemic, individual U.S. states deployed contact tracing apps. The objective is to gather temporal, state-level data on the number of downloads for each app, in order to assess adoption rates and geographical variation.” The user receives five U.S. state/app name pairs for which to collect data and a target date range (2020–2022).

The user interface provides step-by-step guidance with one-sentence explanations for each functionality. The workflow proceeds as follows:

#### Query generation

The user enters the dataset description “Number of downloads over time of contact tracing apps in different U.S. states,” specifies the placeholder name state_and_app_name, and provides the state/app name pairs. With the default of three query templates, the system generates the following:

###### Edited Query Templates

{state_and_app_name} covid app downloads

{state_and_app_name} contact tracing app number of downloads

{state_and_app_name} covid exposure notification app number of downloads

#### Web page retrieval

The user selects U.S. geolocation, sets the date range, enables time-chunking, chooses to retain five results pages per query and a re-ranking threshold of 0.2, retrieving 816 web pages, 153 of which pass the relevance threshold.

#### Data extraction

The system generates the schema:

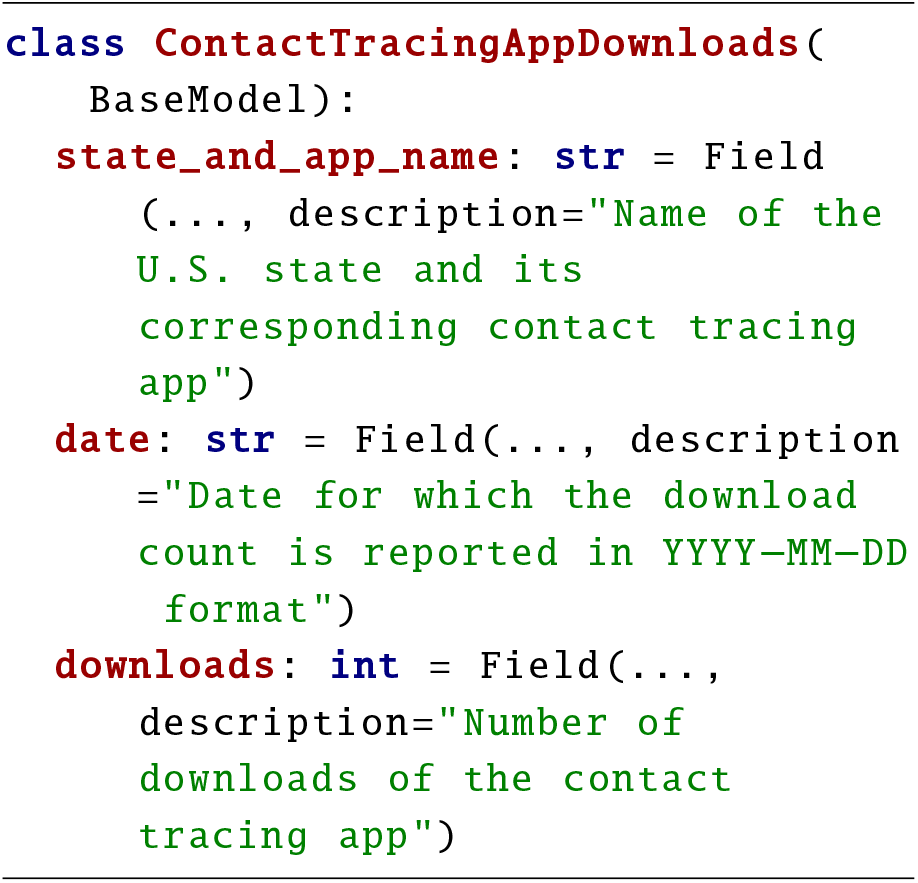

which the user revises to only include the state in the first field, since the corresponding app name can be easily inferred.

The user launches data extraction without specifying additional instructions to the framework, yielding 74 candidate data points.

#### Quality control and cleaning

After automated quality control, the user manual reviews flagged entries. They remove duplicate download counts and address issues such as missing numbers (sometimes reported as percentages rather than raw counts, requiring manual conversion) and missing dates from YouTube sources (when the LLM cannot extract the collapsed video description). The final dataset contains 27 validated data points; Figure 3 shows the time series for the five states.

**Figure 3:**
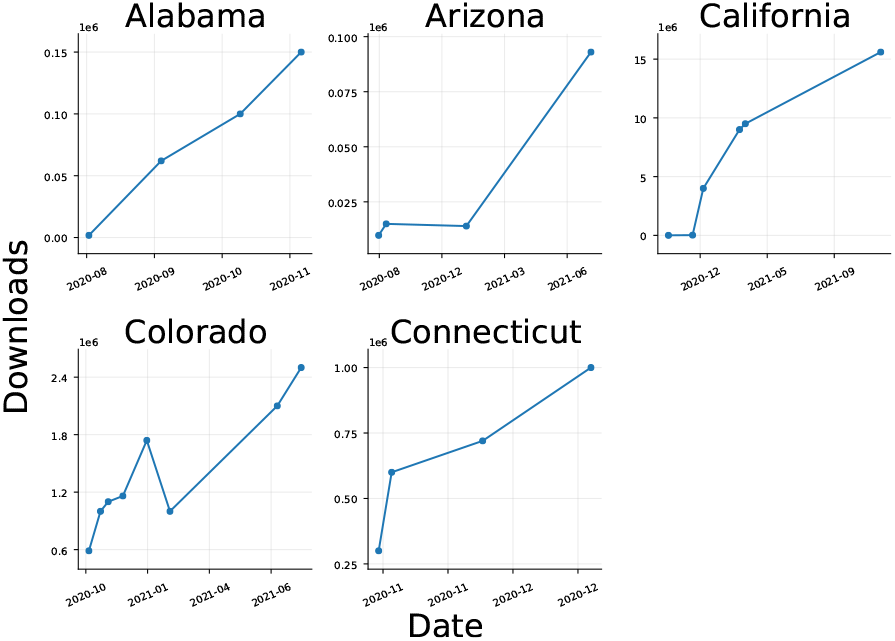
Time series of validated download counts for contact tracing apps in five U.S. states, as collected by the user during the case study.

When compared to the reference dataset, the user’s dataset achieves a recall of 61.4, precision of 84.4, and F1-score of 71.1, slightly below our own experiments. These differences are due primarily to a higher re-ranking threshold, and some manual errors during cleaning.

#### User feedback

The user finds the framework useful and mostly intuitive. Query generation took about 1 minute, web page retrieval 4 minutes (waiting for Google scraping), data extraction 12 minutes (waiting for Crawl4AI extraction), and manual data cleaning 31 minutes after automated quality control. They report that templated queries greatly accelerated repetitive searches. They also appreciate that the framework uncovers “bonus” data for states not in the original five.

Limitations identified include difficulty choosing appropriate parameters (especially the re-ranking threshold) without prior experience, the need to manually verify many entries, frequent duplicates, and some spurious or unclear quality control flags. These issues suggest several improvements: enhancing the framework’s reasoning and acting capabilities (e.g., auto-converting percentages, interacting with web interfaces to reveal hidden data), duplicate detection, and more accurate quality control via more powerful models (e.g., OpenAI o3).

#### Summary

This case study shows that our framework substantially reduces the time and effort needed for web data collection and produces datasets useful for research. However, fewer valid data points were collected than in our own experiments, underscoring a learning curve and the importance of sensitive defaults and a user-friendly interface to ensure effective use by all.

## 7 Conclusion

This paper presents a framework to automate the creation of high-quality research datasets via large-scale web data collection. By combining LLMs with a human-in-the-loop, our method enables efficient extraction of structured data while mitigating various biases that can affect data collection.

Our experimental results and user-centered case study highlight several key strengths of the framework. The template-based search query system, along with efficient re-ranking and filtering methods, make it feasible to collect data at scale across diverse domains. Grounding, bias mitigation, and quality control steps help ensure the integrity and transparency of the resulting datasets—attributes that are crucial for scientific research.

### Limitations

Our framework has several limitations related to both data access and processing capabilities. First, our web data coverage is incomplete: we cannot capture content from sites with access restrictions such as paywalls. Unlike platforms such as Media Cloud, which continuously ingests and archives website data, our framework is also sensitive to web pages being deleted or modified over time—a limitation that impacts both data quality and study reproducibility. To address this, we plan to integrate with the Wayback Machine^7^ to retrieve historical versions of web pages and improve the stability and completeness of collected datasets. Although our framework aims to capture as many relevant data points as possible, it may still miss information due to search engine limitations or filtering steps. As a result, we recommend supplementing its output with manual searches in applications where maximizing coverage is critical. Additionally, while we implement corrections for search engine biases such as recency and geographical bias, our reliance on Google Search still impacts reproducibility due to inherent personalization biases that cannot be fully mitigated.

Currently, the framework’s data processing is limited to text and PDF documents. However, online multimedia content—such as images and videos—also represent valuable research data. Extending support for such content is an important direction for future work.

Finally, the evaluation of our framework could be made more rigorous. Automating the matching of collected data against the reference dataset would make the evaluation process more efficient, as it is currently done manually. Conducting a controlled experiment would also allow us to quantify the manual human baseline for data collection and better measure time savings provided by our framework. While we have included a real example of one user using our framework, a more formal user study with multiple participants and tasks would help identify which aspects of the framework most need improvement to enhance user experience.

## Data Availability

All data produced in the present study are available upon reasonable request to the authors.

## Ethical Considerations

The use of LLMs for web scraping raises important ethical and legal questions (Brown et al., 2024). First, web scraping may violate a website’s terms of service, particularly when collecting copyrighted content or large portions of structured data. Our framework is designed to collect only publicly available content–that is, data not behind authentication barriers–and extracts short snippets rather than full documents. However, the legality of web scraping often depends on the downstream use of the data. It is therefore the responsibility of users to ensure that their data collection complies with relevant laws and regulations, particularly in jurisdictions with stricter data protection laws (European Parliament and Council of the European Union (2016); California Department of Justice (2024)).

Second, while our framework does not deliberately collect personally identifiable information, some scraped content may contain sensitive data. Although this work does not address automated detection or anonymization of such information, we recommend that researchers incorporate appropriate safeguards and anonymization procedures.

Finally, our framework respects ethical scraping practices: it manages request rates to avoid overloading servers and does not attempt to bypass security mechanisms such as CAPTCHAs or paywalls.

## A Template Generation Prompt

Below, we provide the prompt used to automate the generation of Google Search queries. Notably, we instruct Claude to generate queries with the aim of finding specific data points rather than complete datasets. Moreover, rather than using the typical phrase “You are an expert…” to frame our request (Xu et al., 2025), we instruct Claude to be straight-forward and uncreative, as we previously noticed that simpler Google Search queries tend to be more effective at identifying relevant web pages.

Generate {num_templates} Google Search query templates for the following dataset description:

{dataset_description}

Important: Focus on creating concise queries that will find individual data points or small pieces of information, NOT complete datasets or comprehensive lists. Each query should aim to find pages that contain just one or a few relevant pieces of information. Make sure to include important keywords in the query.

Each template should be extremely short and straightforward and the first thing you think of. Do not try to be creative but instead mostly reuse words from the dataset description.

The templates should contain the following placeholder variables, surrounded by curly braces:

{placeholder_names}

Return ONLY a JSON array of template strings. Do not include any explanation or additional text.

## B Schema Generation Prompt

Generate a Pydantic schema based on a list of fields and a data description.

Guidelines:

1. Create one field for each item mentioned in the list of fields and data description.
2. Pick an appropriate data type for each field: int, str or bool. No other types are allowed.
3. Include a description for each field.
4. If there is a date field, use the format YYYY-MM-DD.

The schema should be in the following format:

“‘python

class DataModel(BaseModel):

field_name: data_type = Field(…, description=“Description of the field”)

\# Add more fields as necessary “‘

Example

For the list of fields “country”, “date” and data description “Number of cholera cases”, the output should be:

“‘python

class CholeraCases(BaseModel):

country: str = Field(…, description=“Name of the country for which the cholera case count is reported”)

date: str = Field(…, description=“Date for which the cholera case count is reported in YYYY-MM-DD format”)

cholera_cases: int = Field(…, description=“Number of reported cholera cases”) “‘

Please generate the Pydantic schema for the given list of fields: {schema_fields} and data description: {data_description}.

Return ONLY the schema. Do not include any explanation or additional text.

## C Quality Control Prompt

Below is a dataset collected by an LLM from the web from the prompt:

{dataset_description}

{dataset_csv}

Your task is to examine each row and sanity check it, finding as many potential problems with it as possible and making sure it is consistent with the rest of the data. Output EXACTLY one line per index. Do not miss any rows, do not output any extra rows and do not combine multiple rows’ issues.

Format your response as follows:

index: sentence describing potential problems with the row, or “NA” if you find no issues in a row.

…

## D Baseline Evaluation Prompt

Find as many datapoints as possible from the web for the following dataset description: {dataset_description}. For each datapoint, indicate the date and {list of place-holder names and fields to extract}.

\# For each placeholder name:

The {placeholder_name} in question are:

{list of placeholder values}

…

## E Parameter Settings

**Table 4:**
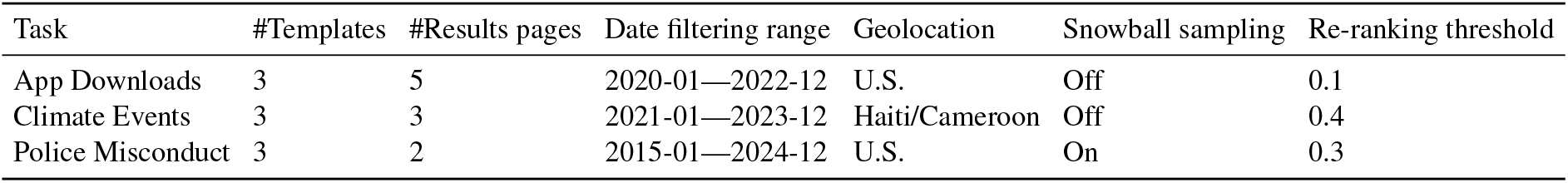
Summary of parameter settings used when evaluating our framework on each data collection task. “#Templates” is the number of generated search query templates. “#Results pages” is the number of Google search results pages scraped per query. “Re-ranking threshold” is the minimum relevance score required to retain a web page during search result filtering. All three were set to limit the number of processed web pages due to computing constraints. “Date filtering range” is the date interval applied to search results. “Geolocation” indicates the country code used as the gl parameter for mitigating geographical bias in Google Search.

| Task | #Templates | #Results pages | Date filtering range | Geolocation | Snowball sampling | Re-ranking threshold |
| --- | --- | --- | --- | --- | --- | --- |
| App Downloads | 3 | 5 | 2020-01—2022-12 | U.S. | Off | 0.1 |
| Climate Events | 3 | 3 | 2021-01—2023-12 | Haiti/Cameroon | Off | 0.4 |
| Police Misconduct | 3 | 2 | 2015-01—2024-12 | U.S. | On | 0.3 |

## Footnotes

1 https://news.google.com/

2 https://lexisnexis.com

3 https://fred.stlouisfed.org/

4 https://www.perplexity.ai/

5 https://openai.com/index/introducing-chatgpt-search/

6 https://github.com/unclecode/crawl4ai, Apache 2.0 license

7 https://web.archive.org/

